# Non-linear Mendelian randomization of vitamin D and C-Reactive Protein: an interrogation of methods

**DOI:** 10.1101/2025.09.10.25335520

**Authors:** Genevieve M Leyden, Fergus W Hamilton, Eleanor Sanderson, George Davey Smith

## Abstract

Mendelian randomization (MR) is an established epidemiological technique which uses genetic variants to strengthen causal inference regarding modifiable exposures. Non-linear MR is an extension to MR which aims to estimate whether the effect differs across the level of the exposure. Many applications of non-linear MR have focused on Vitamin D as an exposure. Using this technique, the study sample is divided into strata, and separate estimates are calculated in each stratum to estimate causal effects at different levels of the exposure (e.g. Vitamin D). For example, a recent study which applied this method identified an apparent protective effect of Vitamin D on C-reactive protein (CRP) levels for those with poor Vitamin D status. However, recent work has highlighted that the commonly used non-linear MR approaches are susceptible to serious bias, suggesting that further methodological development incorporating extensive simulation and empirical investigation is required. In this paper, we provide a commentary on the sources of bias in non-linear MR methods with a re-examination of the relationship between Vitamin D and CRP as an applied example. We highlight the role of negative controls and non-collider variable-based stratification as potential sensitivity tests to identify potential bias for putative non-linear associations in empirical settings.

## Introduction

Mendelian randomization (MR) is an established epidemiological technique which uses genetic variants to strengthen causal inference regarding modifiable exposures in the presence of unmeasured confounding (1, 2). MR analyses test the overall exposure-outcome relationship in the population by incorporating genetic variants found to reliably associate with the exposure into an instrumental variable (IV) analysis (2). Non-linear MR is an extension to MR which aims to estimate whether the effect differs across different levels of the exposure. In non-linear MR analyses the study sample is divided into strata, and separate IV estimates are calculated in each stratum to estimate causal effects across levels of the exposure (3, 4). This approach has the potential to answer important questions around the shape of an exposure-outcome relationship. However recent work has highlighted that the current analytical framework is highly susceptible to bias, suggesting there should be a pause in publication until these issues are addressed (5, 6).

The first developed non-linear MR method is known as the “residual” method, the stratification procedure for which is based on the residuals of the regression of the IV on the exposure (3, 4), thus representing the “IV-free” exposure. MR analyses are then run within each stratum and a non-linear curve is estimated by parametric methods. This strategy aims to avoid collider bias which would be induced by stratifying on the exposure directly (7). The method is dependent on the strict assumption that the genetic effect on the exposure is constant across levels of the exposure. This method has been widely applied (8-14), however results produced by the residual method have been shown to be misleading, owing in part to violation of the required assumption that genetic effects are constant across different levels of the exposure (15, 16). For example, the residual method was applied to investigate putative non-linear effects of Vitamin D on coronary heart disease and all-cause mortality (14). The results of this investigation attributed a protective effect to Vitamin D on all-cause mortality in individuals with low Vitamin D status. Violations of the assumptions of MR are evidenced by a precisely estimated null effect in the overall relationship, but the results of non-linear MR suggesting directionally protective effects of elevating Vitamin D across all strata with substantial benefit in the lowest strata (14). The publication has now been formally retracted (17), due to clearly spurious findings (18, 19) and with the authors stating that their original results were “a logical impossibility” (20). A second paper sharing a similar research question and data as the previous example, describes the relationship between Vitamin D and cardiovascular disease (CVD) risk using the residual non-linear MR method (21). This analysis produced similarly spurious results, where effects in the protective direction were reported for increasing Vitamin D across all strata on CVD risk, but a null effect in the overall population and the study was subsequently retracted (17, 21, 22).

The developers of the original method have since proposed the alternative “doubly-ranked” method which aims to more robustly estimate non-linear exposure-outcome relationships (23). The doubly-ranked method uses a more complex stratification procedure to derive strata with different average levels of the exposure independent of the genetic IV. This is achieved via two-steps. First the population is ranked and assigned into “pre-strata” based on level of the genetic IV. Then within pre-stratum, individuals with the lowest level of the exposure are assigned to stratum-1, and individuals with the next lowest level of the exposure are placed in stratum-2, and so on. This approach relies on the “rank preservation assumption”, i.e. that the relative ranking of individuals by their exposure would be preserved across levels of the genetic IV. This is a strictly weaker parametric assumption regarding the IV and exposure compared to the residual method, with the stratification process being described in more detail in the paper introducing the method (23). While this has been shown in simulations to be less sensitive to heterogeneity in the genetic effect across strata (23), the performance of this methodology has been shown to be highly sensitive to other sources of bias (5, 6, 16).

In this paper we comprehensively interrogate methods that can detect bias in non-linear MR analyses (5, 6). We demonstrate empirically in a re-examination of the relationship between serum 25-hydroxyvitamin D concentration (i.e. Vitamin D) and C-reactive protein (CRP) as described by Zhou et al (9) how sensitivity tests can be applied to detect potential sources of bias in the non-linear association. Further, we demonstrate a stratification approach based on a non-collider variable (Townsend deprivation index) as a powerful sensitivity test for the detection of true non-linear associations. This is based on stratification by a variable which is not itself plausibly influenced by the exposure and outcome, but where sub-groups exhibit different average levels of the exposure. Finally, we advocate that until there is a more reliable demonstration of a non-linear method which is robust to bias, publications applying this method should be paused, or at the very least be viewed with considerable caution.

## Methods

### UK Biobank cohort

UK Biobank is a population-based health research resource consisting of approximately 500,000 people, aged between 38 years and 73 years, who were recruited between the years 2006 and 2010 from across the UK (24). Particularly focused on identifying determinants of human diseases in middle-aged and older individuals, participants provided a range of information (such as demographics, health status, lifestyle measures, cognitive testing, personality self-report, and physical and mental health measures) via questionnaires and interviews; anthropometric measures, BP readings and samples of blood, urine and saliva were also taken (data available at ww.ukbiobank.ac.uk). A full description of the study design, participants and quality control (QC) methods have been described in detail previously (25).

We restricted the sample to individuals of white British ancestry who self-report as “White British” and who have very similar ancestral backgrounds according to the PCA (n=409,703), as described by Bycroft (25). All analyses were conducted on our in-house quality-controlled UKB dataset, with recommended exclusions applied (e.g. genetic outliers, individuals with a high degree of relatedness, minimal relatedness and individuals who have withdrawn consent were excluded from the sample). A full description of all QC applied to the data has been described in detail previously (26). Following exclusions, the total sample size used in downstream analyses was N= 336, 839 individuals of White British ancestry with measured phenotypic data for selected variables. UK Biobank received ethical approval from the Research Ethics Committee (REC reference for UK Biobank is 11/NW/0382).

### Phenotypic measures

Vitamin D (nmol/L) blood biochemistry assays were conducted in the UKB by chemiluminescent immunoassay on the DiaSorin LIAISON XL analyzer and provided in data-field: 30890. C-reactive protein (CRP) (mg/L) blood biochemistry measures were assayed on a Beckman Coulter AU5800 analyzer and provided in data-field: 30710. The distribution of CRP measures in the data is highly skewed and as such values were log-transformed prior to inclusion in analyses. Townsend deprivation index (TDI) in the UKB was calculated immediately prior to participant joining and is based on the preceding national census output areas. Individual participants were assigned a TDI score corresponding to their postcode at recruitment (provided in data-field: 22189), with higher scores corresponding to higher deprivation.

Additional phenotypes were extracted to examine evidence for an interaction effect between an individual’s general health status and the IV-exposure relationship. This was achieved by generating an illness summary score comprising the following traits: Cystatin C (data-field: 30720), forced expiratory volume in one second (FEV1) (data-field: 3063), Albumin (data-field: 30600), cholesterol (data-field: 30690), HbA1C (data-field: 30750) and history of cancer (self-reported diagnoses by doctor) (data-field: 2453). All phenotypic data was accessed via the UKB Research Access Platform (RAP) under UKB application id: 81499.

### Instrumental variables

The vitamin D instrumental variable was based on a focused polygenic risk score (PRS). Use of the focused PRS for Vitamin D is advantageous in that it minimises the potential for bias due to horizontal pleiotropy which may be introduced by including variants which less directly influence variation in Vitamin D. This score comprises 21 genetic variants identified via a conditional association analysis with vitamin D concentration in the UKB at four genetic loci functionally related to vitamin D status: *GC*, *DHCR7*, *CYP2R1* and *CYP24A1* (9, 14, 27). 3 of the 21 variants are rare (minor allele frequency<0.005 in 1000G) and were not present in the UKB, as such the score was generated using the remaining 18 SNPs (9, 14). The weights of scores are provided in **Table S1**) (9, 20). Weighted PRS’ were constructed for individuals in the UKB of White British ancestry using PLINK v2.0 (www.cog-genomics.org/plink/2.0/)(28)).

### Mendelian randomization analyses

Overall exposure-outcome relationships were estimated by standard MR analyses in an individual level setting by two-stage least squares (2SLS) regression using the ‘*ivreg’* R package (29). Covariates included sex, age at assessment, assessment centre, month of assessment, fasting time, genotype chip and the first 10 genetic principal components. Non-linear MR analyses were conducted sing the ‘*SUMnlmr’* R package using the standard settings and Gaussian distribution for continuous outcomes. We selected to use 10 strata for analyses. Non-linear MR analyses using both the “residual” and “ranked” methods were conducted separately.

### Investigations of variables which modify the IV-exposure relationship

A contributing factor to bias in non-linear MR analysis is the potential correlation between the IV and unmeasured confounders affecting the outcome (30). For example, considering Vitamin D as the exposure, an individual’s general health status may plausibly interact with the effect of the genetic IV on measured levels of the exposure. A test for evidence of an interaction effect on the IV-exposure relationship was previously demonstrated by Hamilton et al (6), using a proxy measure for “ill-health”. In that study a composite measure of ill-health was defined using a selection of representative health metrics which included CRP, cystatin-C, albumin, FEV1 and history of dementia, liver disease or cancer prior to recruitment. A similar demonstration is conducted herein, investigating whether the effect of the genetic IV for Vitamin D may be modified by “ill-health”. We based our analysis on a similar selection of health metrics (6), though varied this slightly avoiding the inclusion of CRP in the composite measure.

This was achieved by deriving a “illness summary score” to proxy ill-health status in the sample. Individual participants were assigned scores of 0 or 1 in each of 7 common indicators of ill-health: cystatin C, cholesterol, TDI, HbA1c, FEV1, albumin, and history of cancer. A score of 1 was assigned where measured levels of CRP, cystatin C, cholesterol, TDI, and glycated haemoglobin (HbA1c) resided above the 80^th^ quartile, where measured levels of FEV1 and albumin resided below the 20^th^ quartile and where individuals responded “yes” to a self-reported history of cancer questionnaire. An additive score was then calculated encompassing each of the indicators for ill-health, with the most ill-individuals having a maximum score of 7 and the healthiest having a score of 0. Evidence for an interaction between the illness summary score and the Vitamin D PRS in this setting would indicate that the effect of the IV on an outcome varies depending on the level of the interaction phenotype (i.e. ill-health). Evidence of an effect modification was assessed by linear regression of the measured Vitamin D on the Vitamin D PRS (IV), including an interaction term for the illness summary score.

### MR in participants stratified by a non-collider

We selected TDI as an example of a non-collider because Vitamin D is not expected to influence geographical location or TDI. Further, differences in mean Vitamin D are observed across socio-economic class as indicated by TDI in the UKB. The UKB sample were stratified according to quartiles of TDI, and by season (Summer: April-September; Winter: October-March). Detail of all sample sizes, mean Vitamin D measures and proportion of Vitamin D deficiency across these groupings is provided (**Table S2**). Individual level MR analyses of the relationship between Vitamin D and CRP were performed for each group using the ‘*ivreg’* R package as described above. Further sensitivity tests included individual level MR analysis of Vitamin D on TDI in the overall sample, and both non-linear MR methods.

Secondly, we performed an additional analysis of the effect of Vitamin D on CRP in participants in the UKB stratified by recruitment centre and by season. In this analysis we followed the strategy presented in a recent preprint by Burgess et al (31). The UKB enrolled participants across 22 assessment centres. Participants who attended Stockport and Wrexham assessment centres were dropped due to very low sample sizes (<500 participants). Similarly, participants who attended the Swansea assessment centre in Winter were dropped due to low sample size (<350 participants). As before, individual level MR analyses of the relationship between Vitamin D and CRP were performed for each group using the ‘*ivreg’* R package with adjustment for age, sex, month of assessment fasting time and the first 10 genetic principal components. Sub-group derived MR estimates tested for heterogeneity using Cochran’s Q. Evidence for heterogeneity or non-linearity in the effect was further assessed by fixed effects meta-regression using the ‘*Metafor’* package in R.

## Results

### Estimation of the causal effect of Vitamin D on CRP by MR

We first estimate the causal effect of Vitamin D on CRP in an individual level setting, incorporating a focused PRS for Vitamin D as the genetic IV for Vitamin D. The variance in the trait explained by the focused Vitamin D PRS in the present analysis was estimated at ∼3.9%, aligning with previous studies (14). A precise null effect was observed in the MR analysis of the overall effect of Vitamin D on CRP (Beta= -0.0002, standard error (SE)= 0.0005, P=0.72). The F-statistic for this analysis was large (F=13,727) indicating the result is not likely to be influenced by weak instrument bias. Further detail of the results provided in **Figure 1** and **Table S3**.

**Figure 1.**
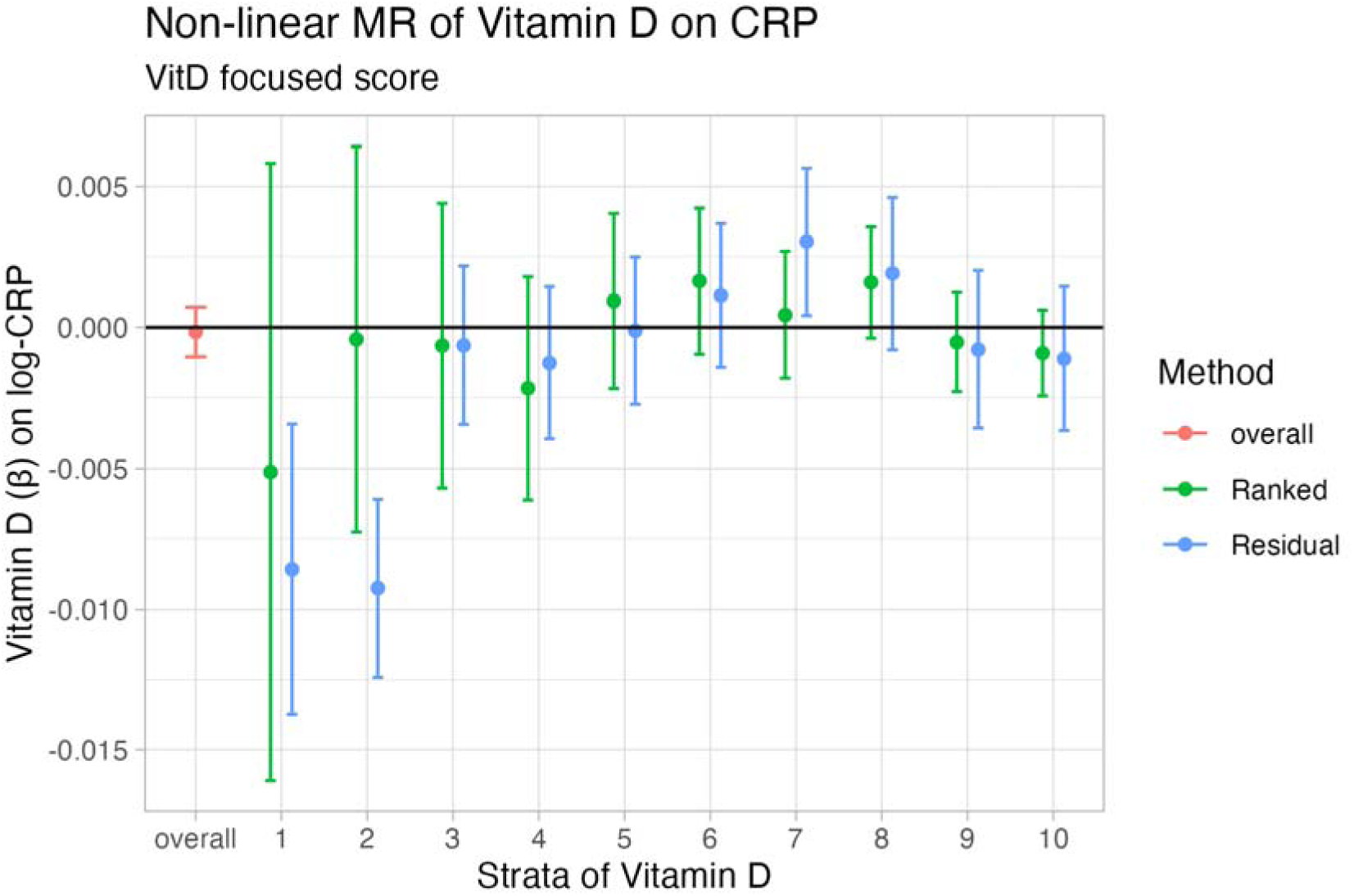
Summary of MR and Non-linear MR estimates of Vitamin D on CRP. The overall estimate derived by standard MR analysis is provided in (red). Results of non-linear MR analyses across 10 strata using the residual and doubly-ranked methods are depicted in blue and green respectively. l

### Estimation of the causal effect of Vitamin D on CRP by non-linear MR

We next examined the Vitamin D-CRP relationship using the residual and doubly-ranked non-linear MR methods. In contrast to the null effect observed when estimating the effect of Vitamin D on CRP in the overall population, the non-linear MR results derived using the residual method indicated evidence of a causal effect in strata of the population with the lowest vitamin D (E.g. Vitamin D PRS stratum 1 (of 10): Beta= -0.008; SE=0.003, P=0.001, stratum 2 (of 10): Beta= -0.004, SE= 0.002, P=9.49×10^-09^). This result recapitulates the findings described by Zhou et al (9) where a non-linear association using the residual method has been described in more detail. Genetically proxied Vitamin D has since been shown to severely violate the “constant genetic effect” assumption which underlies the residual method (15). For this reason, we repeated the non-linear MR analysis using the “doubly-ranked” approach. Our results using the doubly-ranked method support a null effect for Vitamin D on CRP across strata of the exposure, while inconsistent effects observed between strata suggest that performance of the method is likely still sensitive to other bias. Full detailed results are provided **in Figure 1** and **Table S3**.

More detailed evaluations of the performance of the doubly-ranked method in different scenarios have led to caution over its use unless accompanied by sensitivity analyses including triangulation (5, 16) or the incorporation of negative control outcomes, such as sex and age as demonstrated previously in Hamilton et al 2024 (5). In this example, non-null and stratum specific effect estimates were observed in the estimation of Vitamin D on age and sex using both the residual and doubly-ranked non-linear MR methods, where the expected result is null. This negative control assessment of Vitamin D in this non-linear MR setting indicates that bias is introduced in the estimation of stratum specific effects, though it does not indicate the source of bias.

### The Vitamin D IV-exposure relationship is modified by health status

Genetic effect heterogeneity and effect modification of the IV-exposure relationship together are all that is required to generate bias in methods for conducting non-linear MR as presented originally by Small (30) and expanded upon by Hamilton (6). While unmeasured confounding of the exposure and outcome can be assumed for nearly all practical applications of MR, heterogeneity in the genetic effect via any GxE or GxG effect modification may thus induce bias, with the magnitude of the bias being relative to the size of the interaction (6). A variable which could plausibly interact with the genotype effect on an exposure is a person’s general health status, or ‘ill-health’. However, ill-health in population studies is challenging to define and not possible to reliably measure and adjust for. If the genotype effect on the exposure varies by ill-health, it follows that bias in stratified non-linear MR estimates is likely (6).

To demonstrate this empirically, we have investigated evidence of a modification effect by general health status on the IV-Vitamin D relationship using an approximated measure of ill health, referred to as an illness summary score (see methods for detail). Our findings support evidence of an interaction on the genetic IV-exposure relationship for vitamin D, with evidence for an illness effect on Vitamin D (Beta=-0.043, SE=0.001, P<1×10^-300^) and an interaction between illness and the IV (Beta=0.4, SE=0.135, P=3.05×10^-03^). These results align with the interaction effects dissected in further detail by Hamilton et al (6), and help to elucidate potential sources of bias in the stratification procedures used by both the residual and doubly-ranked non-linear MR methods.

### MR in participants stratified by a non-collider variable

The identification of a robust non-linear effect may be aided in a setting where sub-groups with different average levels of the exposure can be generated by stratification of the sample by a non-collider variable (32). To further examine whether the non-linear association between Vitamin D and CRP detected using the residual method may be biased, we explored whether the result is robust to stratification on a non-collider. Individuals in the UKB were assigned a TDI score based on their postcode at recruitment. In the UKB, differences in mean Vitamin D are observed across socio-economic class as indicated by Townsend-deprivation Index (TDI) (8), with differences in the prevalence of Vitamin D deficiency evident between the highest and lowest TDI-Quartile in Summer and Winter months (**Table S2**). As measured Vitamin D is not expected to influence geographical location or TDI, collider bias is unlikely to be an issue. Evidence for an effect of Vitamin D on CRP in the most deficient group would support evidence for a non-linear relationship.

To test this, we have conducted standard individual level MR analyses investigating the relationship between Vitamin D and CRP in strata of the UKB split by TDI-quartile and by season (Summer and Winter). The sample sizes of strata resulting from this stratification procedure ranged from N=38,757:44,818. The results of this analysis produced null effects in all strata (**Figure 2**) (full detailed results **Table S4**). Therefore this finding does not support an effect of Vitamin D on CRP in strata with the lowest Vitamin-D status as described using the residual non-linear MR method (9). As demonstrated in Hamilton 2024, as a negative control we further tested the relationship between Vitamin D on sex and age in each stratum (i.e. outcomes which could not be caused by Vitamin D). The negative control analyses produced null estimates in all strata of TDI (detailed results provided in **Table S4**), indicating that bias was unlikely to have been introduced by the stratification procedure in this setting. We conducted further sensitivity tests of the overall and non-linear relationship of Vitamin D on TDI itself using both non-linear MR methods. The results of this analysis indicated a null effect for the effect of Vitamin on TDI in the overall sample (Beta=-0.001; SE=0.001, P=0.23). While evidence of a non-linear association was detected using the residual method, this was not replicated using the doubly-ranked method (full results provided in **Table S4**, **Figure S1**).

**Figure 2.**
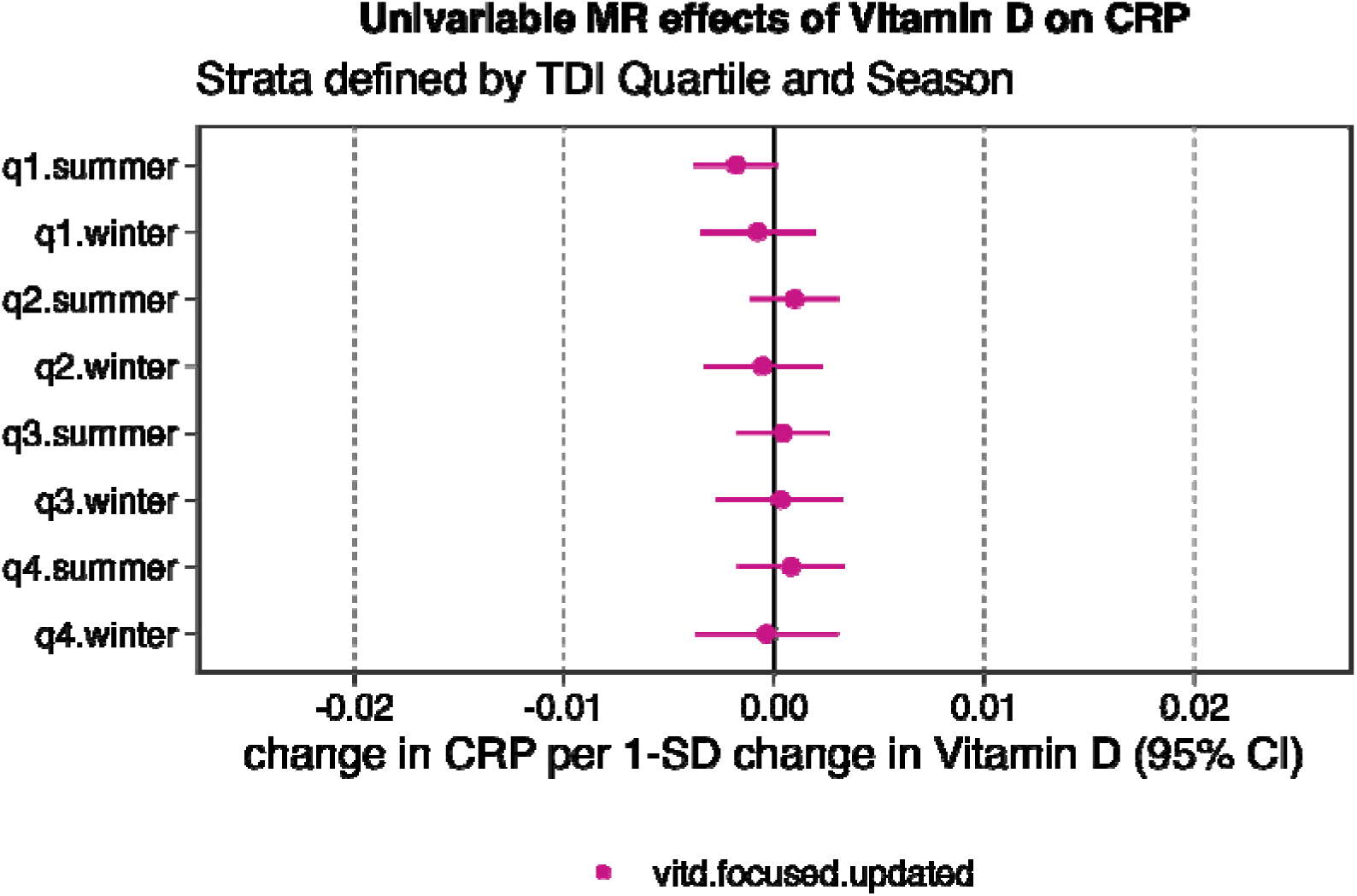
Summary of MR estimates for the effect of Vitamin D on CRP across strata of the UKB population with different average levels of Vitamin D exposure among Townsend deprivation index quartiles and by season.

Lastly, we evaluated the effect of Vitamin D on CRP in individuals in the UKB stratified by recruitment centre using a strategy described in a recent preprint (31). A total of 20 sub-groups could be identified by recruitment centre. Mean Vitamin D values varied from 39.2-56.8 nmol/L across all assessment centres. We note that the power is reduced in this sub-group analysis compared to the stratification by TDI quartiles, with sample sizes varying from N=1,613-31,090 participants. The results of individual level MR analyses conducted within each sub-group were consistent with the null across all groups. Evidence for heterogeneity in the effect estimates was not supported by Cochran’s Q (Q=8.1, degrees of freedom=19, P=0.99) or by meta-regression (Beta=0.0001; SE=0.0001; P=0.57). We repeated this analysis in strata further split by measurements taken between Summer and Winter months (as above). Participants who attended the Swansea clinic during Winter months were omitted from this analysis due to small sample size (N=348), while the sample sizes of remaining groups varied from N=1,259:18,300. Null estimates were observed in all strata in this analysis except for participants measured in Edinburgh in Summer (N=4,599, mean Vitamin D = 41.32nmol/L) where a significant estimate was observed (Beta=-0.01, SE=0.005, P=0.004). However, this result was not replicated in other more highly powered sub-groups with lower mean Vitamin D value, including for example, the sub-group of participants measured in Edinburgh during Winter months (N=7,688, mean Vitamin D = 37.98nmol/L) (Beta=-0.002, SE=0.005, P=0.64). Similarly, evidence for heterogeneity in the effect estimates was not supported by Cochran’s Q (Q=21.5, degrees of freedom=38, P=0.98) or by meta-regression (Beta=0.0001; SE=0.0001; P=0.18). Taken together, evidence for a non-linear association between Vitamin D and CRP is not supported in the present data. A full summary of the subgroups and results from this analysis are provided in **Table S5**.

## Discussion

Non-linear MR is a potentially powerful approach in genetic epidemiology to help gain insight on the shape of exposure-outcome relationships. However, recent work has highlighted outstanding challenges due to bias induced by the stratification procedures used by both the residual and doubly-ranked methods. We examine and summarise via applied demonstrations the detection of bias in non-linear MR methodology which has recently been raised via simulation and empirical analysis (5, 6, 30). In our exploration of potential sources of bias, we decided to focus on the relationship between Vitamin D and CRP as exemplar. The aim of this was to re-examine the non-linear relationship between Vitamin D and CRP which was described using the residual method (9) considering recent developments in the non-linear MR methodology and literature. We have shown by applied example how the evaluation of Vitamin D on CRP by either the residual and doubly-ranked methods is sensitive to bias (as indicated by non-null estimates for Vitamin D on the negative control outcomes age and sex), and evidence of an interaction effect (by ill-health) which could lead to heterogeneous genetic effects in the presence of unmeasured confounding of the exposure-outcome estimation.

Violations of the constant genetic effect and rank preservation assumptions have been demonstrated previously to occur in the presence of unmeasured confounding, with the degree of bias relating to the size of the interaction effect (6). All that is required is an interaction with another variable which can lead to heterogeneity in the genetic effect to induce bias which is unpredictable in direction and effect (6, 15). In our example, strong evidence for an interaction is observed between a proxy-score for ill-health on the IV-Vitamin D relationship which may be contributing to the inconsistent estimates produced via both residual and doubly-ranked methods. However, ill-health is only one of many plausible interacting variables for commonly examined exposures. A primary advantage of conventional MR analysis is to strengthen insight on exposure-outcome relationships in the presence of unmeasured confounding. As such, we highlight the need to address the presence of ubiquitous confounding as an essential area of future development of stratification-based non-linear MR methods.

While the current non-linear MR methodology is heavily reliant on the assumptions underlying the stratification procedure, non-linear associations have been investigated in populations where differences in exposure arise due to a non-collider variable, hence allowing for less strict assumptions about the causal framework. A prominent example of this is highlighted by the evaluation of the effect of alcohol on cardiovascular disease risk factors and events by MR (32-34). This assessment was aided by the presence of genetic variants which reliably predict alcohol intake patterns which are common in East Asian populations, while women who have very low alcohol exposure (due to cultural reasons) can function as a negative control exposure group (35). When considering Vitamin D, we identified Townsend deprivation index (TDI) as a variable where substantial differences in Vitamin D deficiency are observed across quartiles of TDI, which became more prominent when separated by Summer and Winter months. This method of stratification offers a powerful means to sensitivity test putative non-linear relationships within sub-groups with different average levels of the exposure, but which is not itself influenced by the exposure or the IV. Null estimates were observed for the effect of Vitamin D on CRP by standard MR across all TDI strata derived in the present analysis, aligning with the null estimate observed overall. Further, secondary analyses where this was extended to stratification by recruitment centre followed by heterogeneity testing were similarly consistent with the null (31).

The recent progress in the development of non-linear MR methods has highlighted both the importance of and challenges to accurately estimating non-linear exposure-outcome relationships. We advocate that future development of non-linear MR methods should consider the careful selection of controls and, when feasible, non-collider variable-based stratification as a means of sensitivity checking putative bias. In conclusion, we recommend that strategies to mitigate the sources of bias in the current non-linear MR techniques need to be implemented before further applications of the methods.

## Acknowledgements

The authors work in the Medical Research Council Integrative Epidemiology Unit at the University of Bristol, which is supported by the Medical Research Council: MC_UU_00032/1.

## Author Contributions

GML: Writing – primary draft, Methods – formal analysis, code and data acquisition; FWH – Writing – review and editing, Methods – guidance on methodology and code. ES: Writing – review and editing; GDS: Conceptualization, Writing – review and editing.

## Data Availability

All analyses were conducted on individual level UK Biobank data which was accessed via UKB application id: 81499.

